# SARS-CoV-2 RNA and viable virus contamination of hospital emergency department surfaces and association with patient COVID-19 status and aerosol generating procedures

**DOI:** 10.1101/2022.12.22.22283816

**Authors:** Scott C. Roberts, Elliana S. Barbell, Doug Barber, Suzanne E. Dahlberg, Robert Heimer, Karen Jubanyik, Vivek Parwani, Melinda M. Pettigrew, Jason M. Tanner, Andrew Ulrich, Martina Wade, Anne L. Wyllie, Devyn Yolda-Carr, Richard A. Martinello, Windy D. Tanner

**Affiliations:** Department of Internal Medicine, Yale University, New Haven, Connecticut, USA; Department of Epidemiology of Microbial Diseases, Yale School of Public Health, New Haven, Connecticut, USA; Yale Medical School, Yale University, New Haven, Connecticut, USA; Infection Prevention, Yale New Haven Hospital; Department of Emergency Medicine, Yale University, New Haven, Connecticut, USA

## Abstract

**Background:** Infectious aerosols and droplets generated by SARS-CoV-2–positive patient aerosol generating procedures (AGPs), coughing, or exhalation could potentially contaminate surfaces, leading to indirect SARS-CoV-2 spread via fomites. Our objective was to determine SARS-CoV-2 surface contamination frequency in Emergency Department (ED) patient rooms with respect to patient SARS-CoV-2 status and AGP receipt.

**Methods:** Swabs were collected from fixed surfaces or equipment in the rooms of patients under investigation for COVID-19 or known to be SARS-CoV-2-positive. Environmental swabs were tested for SARS-CoV-2 RNA by RT-qPCR; RNA-positive samples were cultured in Vero E6 cells. Room contamination was also evaluated by clinical severity of COVID-19 and time since symptom onset.

**Results:** In total, 202 rooms were sampled: 42 SARS-CoV-2–positive AGP patient rooms, 45 non-AGP SARS-CoV-2–positive patient rooms, and 115 SARS-CoV-2–negative AGP patient rooms. SARS-CoV-2 RNA was detected on 36 (3.6%) surfaces from 29 (14.4%) rooms. RNA contamination was detected more frequently in rooms occupied by non-AGP SARS-CoV-2– positive patients than SARS-CoV-2-positive AGP patients (28.9% vs 14.3%, p=0.078). Infectious virus was cultured from one non-AGP SARS-CoV-2-positive patient room. There was no significant difference in room positivity according to COVID-19 severity or time since symptom onset.

**Conclusion:** SARS-CoV-2 RNA contamination of ED room surfaces was highest and most frequent in rooms occupied by SARS-CoV-2–positive patients who did not undergo an AGP, which may be attributable to disease stage and viral shedding; however, there was no difference in room contamination according to COVID-19 severity or time since symptom onset.

## Introduction

The emergency department (ED) serves as the gateway for hospital admission for most severe cases of COVID-19. Many ED patients with severe COVID-19 require respiratory aid via procedures that may generate aerosols or uncontrolled respiratory secretions. Some procedures are classically considered aerosol generating procedures (AGPs) and include endotracheal intubation for mechanical ventilation, positive pressure ventilation, and cardiopulmonary resuscitation [1]. Other procedures, such as nebulizer therapy and high flow oxygenation, are not clearly AGPs but are frequently used in the care for patients with severe COVID-19; their potential to produce infectious aerosols is unknown.

AGPs are thought to generate higher concentrations of infectious respiratory aerosols than standard coughing or sneezing where aerosols production is negligible. Small particle aerosols may linger in the air for longer periods of time and diffuse farther than larger droplets, increasing the transmission risk from those infected with SARS-CoV-2. Characterizing the extent of SARS-CoV-2 environmental contamination after procedures carries implications for COVID-19 transmission and can help guide infection prevention practices.

We sought to determine the occurrence and viability of SARS-CoV-2 on surfaces of ED rooms occupied by patients suspected of having COVID-19 and whether contamination was associated with COVID-19 severity, time since symptom onset, AGP receipt while receiving care in an ED setting.

## Methods

### Patient identification and room selection

A convenience sample of patients presenting to the Yale New Haven Hospital (YNHH) Adult ED from January to December 2021 were evaluated on arrival for signs and symptoms suggestive of COVID-19. The YNHH Adult ED maintains two campuses that see more than 160,000 patients annually.

Environmental swabs were collected from rooms housing patients under investigation for COVID-19 or known to have an active SARS-CoV-2 infection who underwent AGPs. From April 2021 forward, samples were also collected from rooms of patients known to have an active SARS-CoV-2 infection but who did not undergo an AGP. For this study and in accordance with our institutional policies at the time, AGPs included endotracheal intubation or extubation, manual bag-valve-mask ventilation, cardiopulmonary resuscitation, noninvasive positive-pressure ventilation (NPPV) (both bilevel positive airway pressure [BiPAP] and continuous positive airway pressure [CPAP]), use of high flow oxygenation, bronchoscopy, and nebulizer therapy.

Active SARS-CoV-2 infection was defined as a new positive SARS-CoV-2 test within the 10 days preceding ED presentation, without a prior positive test in the 90 days preceding this test. COVID-19 status was not typically known for AGP patients prior to environmental sampling; as a result, rooms housing AGP patients who ultimately tested negative for COVID-19 were enrolled as negative controls. Patient COVID-19 status at the time of the ED encounter through 14 days following the encounter was verified using the electronic medical record. Nucleic acid amplification testing for SARS-CoV-2 in ED patients was performed on nasopharyngeal or mid-turbinate nasal specimens using U.S. Food and Drug Administration emergency use authorized platforms. YNHH environmental cleaning protocols dictate that accessible surfaces and non-disposable medical equipment should be cleaned and disinfected using a disinfectant effective against SARS-CoV-2 prior to the next occupant. Environmental samples collected from rooms where patients died or were discharged prior to SARS-CoV-2 testing were excluded from the analysis. The study protocol was approved by the Yale University Institutional Review Board (IRB) and the Yale Human Research Protection Program (IRB# 2000029221).

### Environmental surface sample collection

Samples were collected using SANI-MacroSwabs (Sanigen Co., Ltd, Anyang-si, Gyeonggi-do, South Korea) saturated with a viral transport medium containing no agents known to deactivate SARS-CoV-2 [2]. Five samples were collected from each room while occupied by the suspected COVID-positive patient or immediately following patient discharge or transfer, but prior to room cleaning and disinfection. Four samples were from predetermined fixed surfaces, which were selected based on touch frequency and proximity to the aerosol source: high touch surfaces within and further than 6 feet from the patient (bedrail, room door handle, respectively), and low touch surfaces within and further than 6 feet from the patient (vital signs monitor frame, air return vent [procedure light in resuscitation rooms], respectively) (see Supplementary Figure S-1A, Supplementary Table S-1). A fifth sample was taken from the reusable equipment surface associated with the AGP (see Supplementary Figure S-1B): high flow oxygen controller, NPPV machine, video laryngoscope, or mechanical ventilation controller. If nebulizer therapy was administered or the patient did not receive an AGP, the oxygen gauge directly behind the patient was swabbed.

### SARS-CoV-2 RNA and infectious SARS-CoV-2 detection

Environmental swab elution and viral deactivation were conducted under biosafety-level 2+ conditions. Tissue culture work was conducted in the biosafety-level 3 laboratory facilities at Yale University. All laboratory protocols were approved by the Yale University Biological Safety Committee. Samples were stored at 4°C up to 72 hours or frozen at -80°C until processing. Swabs were eluted with 5 mL sterile phosphate buffered saline (PBS) and pulse vortexed for 2 minutes. One milliliter of swab eluate was aliquoted for RNA extraction using the Qiagen RNeasy Mini® RNA extraction kit (Qiagen, Hilden Germany). The remaining 4 mL eluate was frozen at -80°C for tissue culture of samples positive for SARS-CoV-2 RNA by reverse transcriptase quantitative PCR (RT-qPCR). RT-qPCR testing of environmental samples utilized the U.S. Centers for Disease Control (CDC) N1 primer/probe set [3]. The CDC human RNase P (RP) extraction control primers were not utilized because the samples were not directly from humans.

Samples testing positive for SARS-CoV-2 RNA were cultured using the remaining eluate to determine whether infectious virus was present. The eluate was passed through a 0.45 µL syringe filter and concentrated down to 200 µL using an Amicon Ultra-4 30 kDa Ultracel-PL filter (Millipore Sigma, St. Louis, Missouri, United States), then inoculated onto T25 flasks of Vero E6 cells overexpressing TMPRSS2 and ACE2 [4]. An aliquot of culture media was placed in Buffer AVL (lysis buffer) (Qiagen, Hilden, Germany). Flasks were examined at 4 days post infection for cytopathic effect, and a second culture media aliquot was removed and placed in Buffer AVL. Flasks were fixed with 4% paraformaldehyde and stained with a 0.05% crystal violet solution. RNA extraction from AVL buffer aliquots was performed using QIAamp viral RNA Mini kit (Qiagen, Hilden, Germany). RT-qPCR was performed on pre- and post-culture samples within the same assay run to determine if there was a reduction in cycle threshold due to SARS-CoV-2 proliferation.

### COVID-19 status of previous occupant of sampled ED rooms

Bed tracking was performed, and the electronic health record was reviewed to determine the COVID-19 status of the immediately prior room occupant of sampled ED rooms and evaluate whether SARS-CoV-2-contaminated rooms occupied by a COVID-19-negative patient were associated with a prior occupant with COVID-19.

### Relationship between patient symptom presentation and ED surface SARS-CoV-2 contamination

The electronic health record was used to determine time since COVID-19 symptom onset and symptom severity for patients who were SARS-CoV-2-positive at the time of room sampling. A previously utilized COVID-19 ordinal severity index (0 = not hospitalized and no limitations of activities, up to 8; death due to COVID-19) [5] and days from symptom onset to disease presentation were calculated for all patients actively-positive SARS-CoV-2 patients (n = 83).

### Statistical analysis

Differences in SARS-CoV-2 RNA contamination of AGP and non-AGP COVID-positive patient rooms was analyzed using Fisher’s Exact Test. Differences in SARS-CoV-2 RNA concentrations were analyzed using the Kruskal-Wallis test. The relationships between room contamination and COVID-19 severity or time since COVID-19 symptom onset was analyzed by the independent samples median test. IBM SPSS version 28.0 was used for statistical analysis with a p-value of < 0.05 deemed significant.

## Results

### Demographics and patient COVID-19 positivity

A total of 202 rooms housing patients known or suspected of having COVID-19 who received SARS-CoV-2 testing within the 10 days preceding environmental sampling were sampled, generating 1010 environmental swab specimens. Samples were collected from various room types, including resuscitation bays (n = 69, 34.2%), airborne infection isolation rooms (n = 56, 27.7%), and normal ED rooms (n = 77, 38.1%). AGPs were performed during the patients’ occupancies in 157 (77.7%) of sampled rooms. The most frequently performed AGP in the sampled rooms was intubation (n = 52), followed by NPPV (BiPAP [n = 47] and CPAP [n=2]), high flow oxygenation (n = 34), and nebulizer therapy (n = 13). One room housed a patient who received manual bag-valve-mask ventilation and eight rooms housed patients who received multiple AGPs. A total of 87 patients (43.1%) were confirmed positive for SARS-CoV-2 after sampling. Approximately half of the COVID-19-positive patients underwent an AGP (n = 42, 48.3%). Of the 42 COVID-19-positive patients who underwent an AGP, 66.7% received high flow oxygenation (n=28), 9.5% (n=4) each received NPPV, nebulizer treatments, or were intubated, and 4.8% (n=2) received multiple AGPs. Table 2 lists patient COVID and AGP status by AGP type.

### SARS-CoV-2 surface contamination

Thirty-six of the 1010 swabs (3.6% of swabs) from 29 of the 202 rooms (14.4% of rooms) sampled were positive for SARS-CoV-2 RNA (Table 1). Of rooms occupied by patients confirmed as SARS-CoV-2-positive, 19 of 87 rooms (21.8%) were contaminated with SARS-CoV-2 RNA on at least one surface. SARS-CoV-2 RNA was detected more frequently on surfaces in COVID-positive patient rooms when AGPs were not performed (n = 13 out of 45 rooms, 28.9%), than when AGPs did occur (n = 6 out of 42 rooms, 14.3%, p = 0.078), however the difference was not statistically significant. SARS-CoV-2 RNA was detected in 10 of the 115 (8.7%) rooms occupied by COVID-negative patients. In SARS-CoV-2-contaminated rooms occupied by COVID-negative patients, the average surface contamination was 24.1 RNA copies/100 cm^2^. In SARS-CoV-2-contaminated rooms of COVID-19 positive patients who received an AGP, the average surface contamination was 52.1 RNA copies/100 cm^2^. In SARS-CoV-2-contaminated rooms housing COVID-19-positive patients who did not receive an AGP, the average surface contamination was 147.6 RNA copies/100 cm^2^. Surface contamination concentration exhibited the same pattern as surface contamination frequency (i.e., highest in non-AGP COVID-positive patient rooms; lowest in COVID-negative patient rooms); however, the results were not statistically significant (p=0.239).

**Table 1:** Percentage of rooms and surface swabs testing positive for SARS-CoV-2 RNA or infectious virus by RT-qPCR or culture

| COVID-19 status | Aerosol generating procedure (AGP) | Number of rooms sampled | Number of rooms (%) positive for SARS-CoV-2 RNA | Number of swabs collected | Number of swabs (%) positive for SARS-CoV-2 RNA | Average SARS-CoV-2 RNA copies* per 100 cm<sup>2</sup> swabbed surface area | Number of swabs positive by viral tissue culture |
| --- | --- | --- | --- | --- | --- | --- | --- |
| Positive | Yes | 42 | 6 (14.3%) | 210 | 8 (3.8%) | 52.1 | 0 |
| Positive | No | 45 | 13 (28.9%) | 225 | 16 (7.1%) | 147.6 | 1 |
| Negative | Yes | 115 | 10 (8.7%) | 575 | 12 (2.1%) | 24.1 | 0 |
| **Total** |  | **202** | **29 (14.4%)** | **1010** | **36 (3.6%)** |  | **1** |

\*As determined by RT-qPCR curve determined from known quantities of PCR

Of the six SARS-CoV-2 RNA-contaminated rooms of COVID-19-positive patients where AGPs occurred, high flow oxygenation occurred in five rooms, while nebulizer therapy occurred in one. No room surfaces were contaminated with SARS-CoV-2 RNA after patients with active COVID-19 underwent intubation or NPPV. Table 2 summarizes environmental detection of SARS-CoV-2 by AGP type and patient COVID-19 positivity.

**Table 2:** Patient COVID-19 positive and/or room surface positivity by aerosol generating procedure (AGP) type

| **Sample types** | **BiPAP<sup>a</sup>** | **HFNC<sup>b</sup>** | **Intubation** | **Nebs<sup>c</sup>** | **Other<sup>d</sup>** | **Multiple** | **Total**<br>**(n)** |
| --- | --- | --- | --- | --- | --- | --- | --- |
| AGPs performed | 30%<br>(n=47) | 22%<br>(n=34) | 33%<br>(n=52) | 8%<br>(n=13) | 2%<br>(n=3) | 5%<br>(n=8) | 157 |
| AGP & COVID+<br>patient | 10%<br>(n=4) | 67%<br>(n=28) | 10%<br>(n=4) | 10%<br>(n=4) | 0%<br>(n=0) | 5%<br>(n=2) | 42 |
| AGP, COVID+<br>patient, &<br>surface+ | 0%<br>(n=0) | 83%<br>(n=5) | 0%<br>(n=0) | 17%<br>(n=1) | 0%<br>(n=0) | 0%<br>(n=0) | 6 |
| AGP, COVID-<br>patient, &<br>surface+ | 30%<br>(n=3) | 10%<br>(n=1) | 30%<br>(n=3) | 10%<br>(n=1) | 10%<br>(n=1) | 10%<br>(n=1) | 10 |

a. BiPAP: Bilevel positive airway pressure; b. High Flow: High flow nasal cannula; c. Nebs: Nebulizer therapy d. Other = Continuous positive airway pressure, Bag-valve-mask ventilation

SARS-CoV-2 RNA was most frequently detected on air duct return vents (n = 13, 36.1%), followed by bedrails (n = 10, 24.4%), reusable equipment (n = 4, 11.1%), monitors (n = 4, 11.1%), door handles (n = 3, 8.3%), and procedure lights (n=2, 5.6%). Supplementary Table S-2 lists positive room surfaces according to patient COVID and AGP status. SARS-CoV-2 RNA contamination ranged from 5 – 687 copies/100 cm^2^ on vents, 6 – 74 copies/100 cm^2^ on bedrails, 32 – 39 copies/100 cm^2^ on door handles, 16 – 65 copies/100 cm^2^ on reusable equipment, and 22 – 504 copies/100 cm^2^ on the monitor frame (Supplementary Table S-2). Most contaminated patient rooms had only a single SARS-CoV-2-positive surface; however, five rooms had two positive surfaces, and one room had three positive surfaces.

### SARS-CoV-2 viability

One of the 36 samples positive for SARS-CoV-2 RNA was also positive by viral tissue culture, with extensive cytopathic effect observed (Figure 1). The swab was from the bedrails in a non-AGP COVID-19-positive patient room. The original swab C_t_ value was 35.7 (RNA copies = 147.7). The cell culture media from the inoculated tissue culture sample exhibited a C_t_ reduction from > 40 to 13 on days 0 and 4, respectively. Significant reductions in C_t_ or extensive cytopathic effect in tissue culture were not observed for any other SARS-CoV-2 RNA-positive swabs.

**Figure 1.**
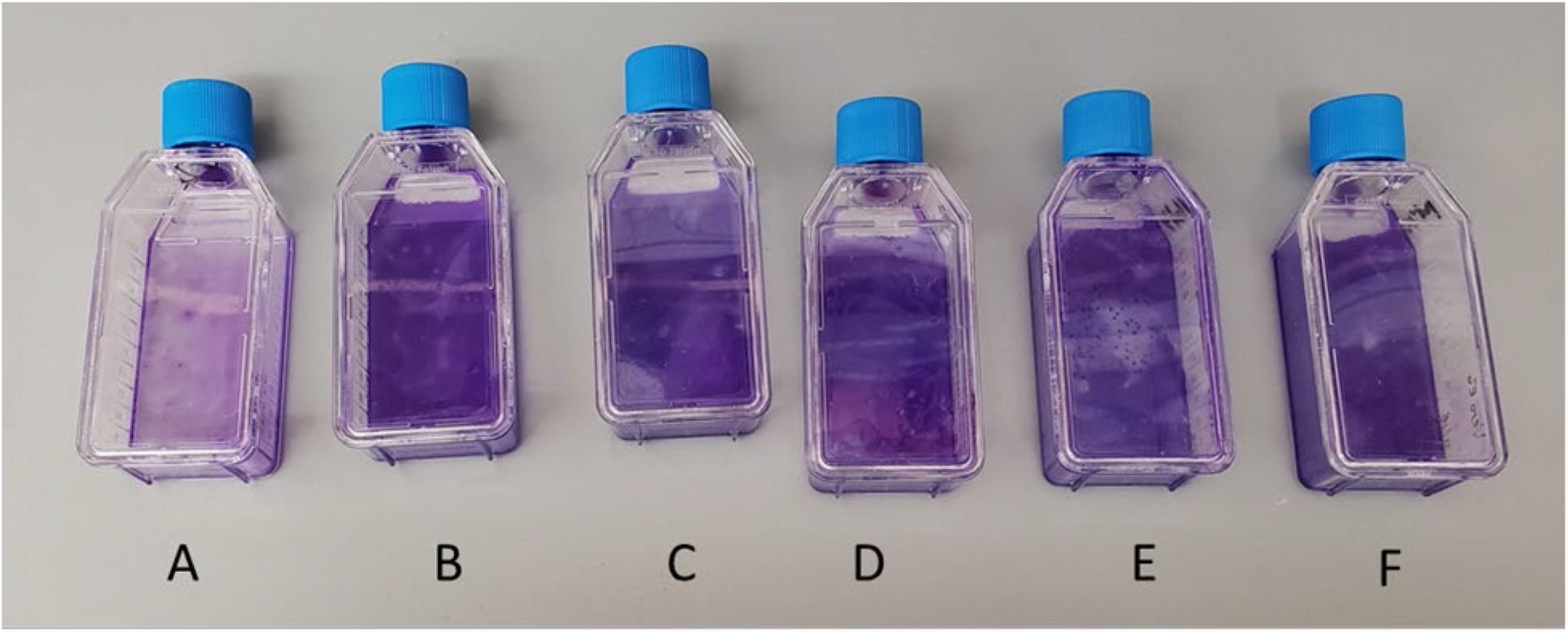
Vero E6 cell culture of environmental samples. Cell culture of SARS-CoV-2 RNA positive Sample A (bedrail) using ACE2+ TMPRSS+ Vero E6 cells exhibited extensive cytopathic effect; B-F are examples of samples that were positive for SARS-CoV-2 RNA but did not exhibit any cytopathic effect in cell culture.

### COVID-19 status of previous occupant of sampled ED rooms

Thirteen of the 202 sampled rooms (6.4%) were occupied by a COVID-19 positive patient immediately preceding the patient occupying the room at the time of sampling. In one of the 13 instances (7.7%), the room surface was positive for SARS-CoV-2 contamination while occupied by a COVID-19 negative patient at the time of sample collection. However, the positive surface was a reusable piece of equipment (high flow apparatus) that was not used during the prior occupancy of the COVID-19 positive patient.

### Relationship between patient symptom presentation and ED surface SARS-CoV-2 contamination

For patient rooms positive for SARS-CoV-2 surface contamination, the median severity for COVID-19 positive patients on presentation was 5 [interquartile range (IQR): 2.5 – 6]; the median number of days from symptom onset to emergency department presentation was 4 days (IQR 3 – 7).

For patient rooms that were negative for SARS-CoV-2 surface contamination, the median severity for COVID-19 positive patients was 6 (IQR 4.75 – 6, p = 0.259 when compared to positive rooms); the median number of days from symptom onset to ED presentation was 7 days (IQR 3 – 8.5, p = 0.507 when compared to positive rooms). Thus, rooms contaminated with SARS-CoV-2 had a higher frequency of housing patients with milder disease earlier in the time course of infection, however this was not statistically significant.

## Discussion

SARS-CoV-2 RNA contamination was detected on at least one surface in over 20% of the sampled rooms housing patients with COVID-19. Surface contamination was detected more frequently and at higher concentrations in rooms of COVID-19 patients who did not have an AGP, as was the one sample with viable virus. We suspect this observation is due to the time course of COVID-19 and reflects the natural disease progression in which viral loads peak in the respiratory tract around the time of symptom onset, even in patients who go on to develop severe disease [6].

Additionally, the primary mechanisms driving severe disease where patients require AGPs later in the disease course are likely immune-mediated inflammation with lowered viral loads [7]. We suspect most COVID-positive AGP patients were suffering from the hyper inflammatory phase of their illness where viral load was diminished, supporting the lower rate of environmental SARS-CoV-2 detection. We observed a trend of a lower proportion of contaminated rooms for patients with more severe disease, supporting this observation. This carries infection control implications; while AGPs may lead to more extensive room contamination with respiratory secretions, a focus on AGPs may not be as essential as mitigating transmission earlier in the disease course when viral transmission potential is greatest, regardless of aerosol deposition.

Of patients with COVID-19 who underwent an AGP, surface contamination occurred most frequently following the use of high flow oxygenation and nebulizer therapy. These therapies are not typically considered AGPs [1, 8] although concern has been raised that these procedures may facilitate human to human transmission of SARS-CoV-2 [9-12]. We did not detect surface contamination following intubation or non-invasive positive pressure ventilation of COVID-19 patients, procedures thought to induce aerosols. The sampled surface areas were small, and dispersed aerosols may also have diminished the sensitivity of our approach to detect SARS-CoV-2 [8].

We observed that 9% of rooms were contaminated with SARS-CoV-2 when patients were COVID-19-negative. In most cases, the air duct return vent was most frequently implicated, suggesting prior contamination. Given the busy turnover of ED rooms, prior patients or staff could have contributed to the air duct return vent contamination, as these surfaces were not routinely disinfected between patients. This also highlights the ability of upward airflow, even in rooms maintained without negative pressure, to move SARS-CoV-2 aerosols to locations that will not result in viral transmission. Retrospective review of SARS-CoV-2 contaminated room occupancy prior to the time of sampling revealed only one case in which a COVID-positive patient occupied the contaminated room immediately prior to its occupancy by a COVID-negative patient at the time of sampling. In this instance, the contaminated surface (reusable equipment) was not present during the prior patient’s stay, emphasizing the risk of SARS-CoV-2 spread via mobile equipment.

Only one sample exhibited cytopathic effect consistent with growth of SARS-CoV-2 in cell culture. This sample was collected from the bedrails in a room housing a COVID-positive non-AGP patient at the time of sampling. Infectious virus has rarely been recovered from hospital surfaces and other reports also indicate that these samples are typically within close range of the patient [13-15]. Viable SARS-CoV-2 has been shown to remain viable on surfaces for as long as 21 days with a half-life of approximately 2-5 days [16], while SARS-CoV-2 RNA only exhibits a 1-log reduction over the same amount of time [17]; therefore, failure to detect viable virus in more samples with higher viral concentrations per 100 cm^2^ is not unexpected and supports the current thought on the minimal role of surface and fomite transmission in spreading SARS-CoV-2, as others have noted [18].

Limitations of this study include the single hospital evaluation, which may limit generalizability, although samples were collected from two campuses. Further studies should confirm these findings in non-ED settings where AGPs are performed, such as operating rooms and intensive care units (ICUs). Other studies have reported a much higher frequency of SARS-CoV-2 RNA detection [19]; however, few studies have reported SARS-CoV-2 in emergency department settings [20], which have significantly higher room turnover rates and a more limited period in which contamination can occur compared to ICUs. Because we selectively sampled rooms where patients were either known to have COVID-19 or those who were under investigation for COVID-19 due to signs and symptoms suggestive of an active infection, these findings do not represent the general prevalence of environmental SARS-CoV-2 RNA contamination in an ED setting. It is unclear if findings would be replicated in an asymptomatic infected patient population. As we retrospectively reviewed the electronic medical record for COVID-19 severity and time since symptom onset, misclassification bias is possible. Lastly, it is possible more samples would have tested positive for SAR-CoV-2 RNA if the entire sample, as opposed to only one-fifth, underwent RNA extraction and PCR.

There remains an incomplete understanding of how the range of specific procedures involving the respiratory tract may increase environmental contamination and the risk of transmission of SARS-CoV-2 and other respiratory pathogens. Future investigation will be necessary to clarify both how the spectrum of respiratory tract procedures/treatments alter the generation of large droplets and small particle aerosols and how these changes may impact viral transmission and environmental contamination.

## Data Availability

All data produced in the present study are available upon reasonable request to the authors.

## Funding

This work was supported by U.S. Centers for Disease Control and Prevention Broad Agency Announcement 75D301-20-R-68024 – Applied Research to Address the Coronavirus (COVID-19) Continued Public Health Emergency – *Developing Strategies for Protection, Prevention, & Control in Communities* [contract# 75D30120C09810].

## Acknowledgements

We are grateful to Dr. Barney Graham at the U.S. National Institutes of Health for sharing his Vero E6 cell line.

We are grateful to our colleagues Alisha Shams (CDC), Laura Rose (CDC), Judith Noble-Wang (CDC), Allison Perry (CDC), Carrie Whitworth (CDC), Nancy Burton (NIOSH), Sujan Reddy (CDC), and Geun Woo Park (CDC) for their guidance on this study.

## Declarations of Conflicts of Interest

The authors declare no competing interest.

## Notes

### Competing Interest Statement

The authors have declared no competing interest.

### Author Declarations

The IRB of Yale University waived ethical approval for this work.

