## Supplementary material for "SARS-CoV-2 RNA and viable virus contamination of hospital emergency department surfaces and association with patient COVID-19 status and aerosol generating procedures": Table S-1

Supplemental data

Table S-1. ED room surfaces positive for SARS-CoV-2 contamination

| **Surface** | **Surface area sampled** | **Number of samples collected** | **Number of positive swabs** |
| --- | --- | --- | --- |
| Bedrail (high touch, <6 feet from patient) | 129 cm^2^ | 202 | 10 |
| Door handle (high touch, >6 feet from patient) | 65 cm^2^ | 202 | 3 |
| Air return vent (low touch, >6 feet from patient) | 155 cm^2^ | 133 | 13 |
| Vital signs monitor frame (low touch, <6 feet from patient) | 194 cm^2^ | 202 | 4 |
| Resuscitation room procedure light (in lieu of air return vents) | 232 cm^2^ | 69 | 2 |
| Equipment: Glidescope frame | 155 cm^2^ | 4 | 0 |
| Equipment: Noninvasive positive pressure ventilation control screen frame (BiPAP; CPAP) | 226 cm^2^ | 53 | 0 |
| Equipment: High flow oxygen control | 219 cm^2^ | 36 | 1 |
| Equipment: Oxygen gauge (swabbed for nebulizer treatments and when no AGP was administered) | 97 cm^2^ | 56 | 1 |
| Mechanical ventilation control screen frame | 226 cm^2^ | 49 | 2 |
