## Supplementary material for "SARS-CoV-2 RNA and viable virus contamination of hospital emergency department surfaces and association with patient COVID-19 status and aerosol generating procedures": Table S-2

Supplemental data

Table S-2. Estimated RNA copies per 100 cm^2^ of swabbed surface area for each positive sample

| **Surface** | **Patient COVID result** | **Patient AGP* status** | **RNA copies per 100 cm^2^** |
| --- | --- | --- | --- |
| Vent | Positive | **No AGP** | 77.8 |
| Vent | Negative | **AGP** | 9.6 |
| Door | Negative | **AGP** | 32.3 |
| Bedrail | Positive | **AGP** | 59.0 |
| Vent | Negative | **AGP** | 9.6 |
| Vent | Positive | **No AGP** | 163.6 |
| Vent | Negative | **AGP** | 7.5 |
| Equipment | Negative | **AGP** | 15.9 |
| Vent | Positive | **No AGP** | 71.7 |
| Vent | Positive | **No AGP** | 686.6 |
| Bedrail | Negative | **AGP** | 18.2 |
| Door | Negative | **AGP** | 39.4 |
| Bedrail | Positive | **AGP** | 73.9 |
| Bedrail | Positive | **No AGP** | 6.4 |
| Procedure light | Negative | **AGP** | 50.1 |
| Equipment | Negative | **AGP** | 46.8 |
| Vent | Positive | **No AGP** | 28.6 |
| Vent | Positive | **No AGP** | 36.6 |
| Bedrail*** | Positive | **No AGP** | 114.5 |
| Monitor | Positive | **No AGP** | 504.7 |
| Vent | Positive | **No AGP** | 5.0 |
| Door | Positive | **No AGP** | 36.1 |
| Vent | Positive | **No AGP** | 14.9 |
| Vent | Positive | **No AGP** | 50.8 |
| Bedrail | Positive | **No AGP** | 12.8 |
| Bedrail | Positive** | **No AGP** | 538.1 |
| Bedrail | Positive | **No AGP** | 13.5 |
| Monitor | Positive | **AGP** | 147.9 |
| Bedrail | Positive | **AGP** | 7.4 |
| Equipment | Positive | **AGP** | 64.8 |
| Equipment | Negative | **AGP** | 25.8 |
| Procedure light | Negative | **AGP** | 11.9 |
| Monitor | Negative | **AGP** | 22.2 |
| Bedrail | Positive | **AGP** | 4.7 |
| Vent | Positive | **AGP** | 22.3 |
| Monitor | Positive | **AGP** | 37.2 |

* AGP: Aerosol-generating procedure; **COVID-19-positive by self-report. Not tested at Yale New Haven Hospital. ***Indicates sample from which viable SARS-CoV-2 virus was recovered.
