## Supplementary figures and images for "SARS-CoV-2 RNA and viable virus contamination of hospital emergency department surfaces and association with patient COVID-19 status and aerosol generating procedures"

### Figure S-1A

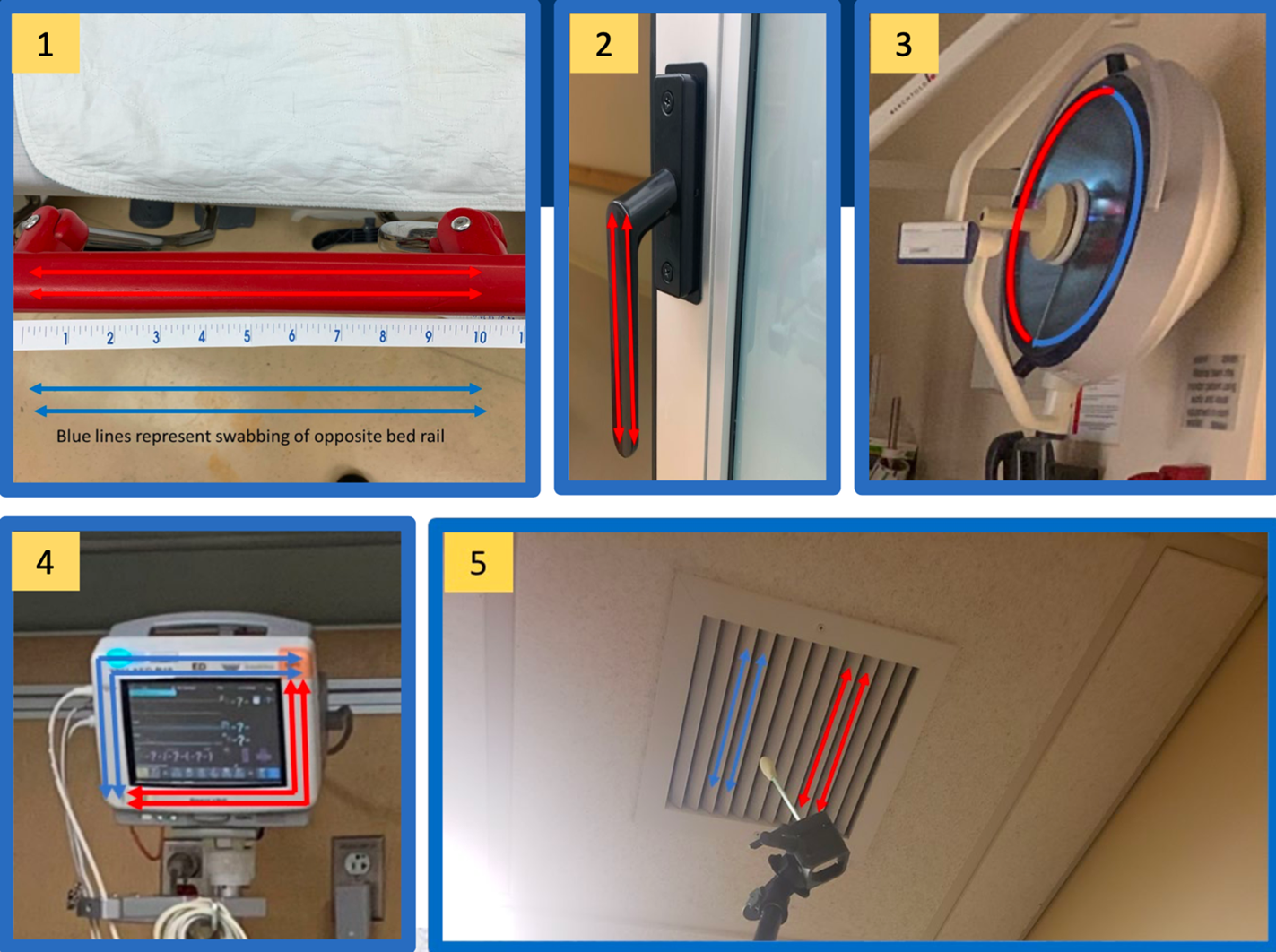

### Figure S-1B

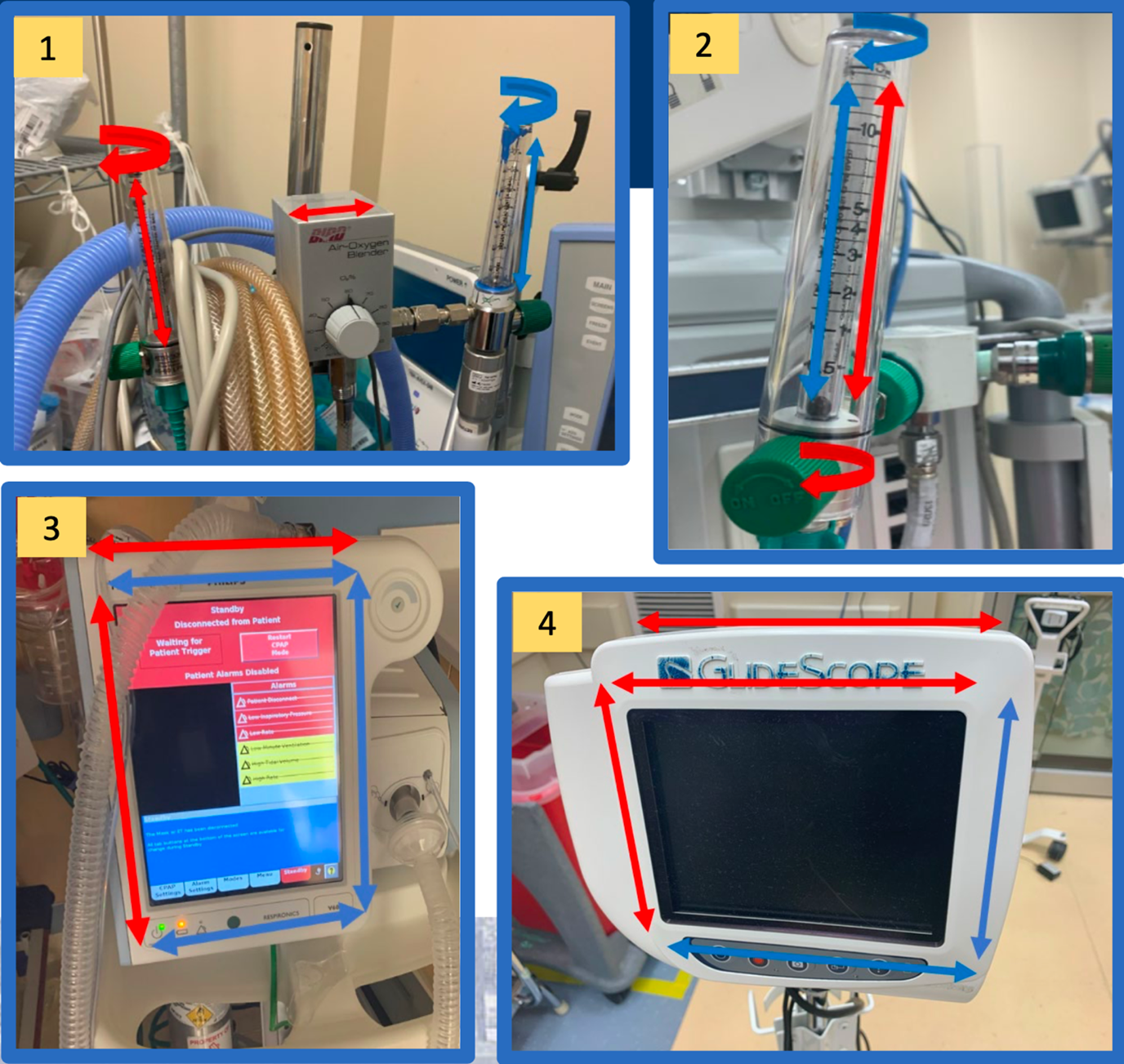
